# Antenatal care policy in high-income countries with a universal health system: a scoping review

**DOI:** 10.1101/2021.09.03.21263024

**Authors:** Andreia Soares Goncalves, Isabel Maria Ferreira, Márcia Pestana-Santos, Christine McCourt, Ana Paula Prata

## Abstract

The availability, effectiveness, and access to antenatal care are directly linked with good maternal and neonatal outcomes, making antenatal care an important determinant in health. But to be effective, care must always be appropriate, not excessive, not insufficient. Perinatal outcomes vary within and between countries, raising questions about practices, the use of best evidence in clinical decisions and the existence of clear and updated guidance.

Through a scoping review methodology, this study aimed to map the available antenatal care policies for low-risk pregnant women in high-income countries with a universal health system founded on the Beveridge Model.

Following searches on the main databases and grey literature, the authors identified and analysed ten antenatal care policies: Australia, Denmark, Finland, Iceland, Italy, Norway, Portugal, Spain, Sweden and the United Kingdom. Some policies were over 10 years old, some recommendations did not present a rationale or context, others were outdated, or were simply different approaches in the absence of strong evidence. Whilst some recommendations were ubiquitous, others differed either in the recommendation provided, the timing, or the frequency. Similarly, we found wide variation in the methods/strategy used to support the recommendations provided. These results confirms that best evidence is not always assimilated into policies and clinical guidance. Further research crossing these differences with perinatal outcomes and evaluation of cost could be valuable to optimise guidance on antenatal care. Similarly, some aspects of care need further rigorous studies to obtain evidence of higher quality to inform recommendations.

## Background

Pregnancy and birth are major life events: for women, for a family, and the society(1). Mother and newborns’ health is paramount for a ‘good start’ in life and without the right care, this ‘start’ can be a stressful, damaging, or even a tragic event(2). Effective maternity care is, therefore, a pivotal global health policy(3), mirrored in the Sustainable Developmental Goals(4) agenda for 2030, and, unsurprisingly, extraordinary attention to antenatal care is paid by the health services all over the world(1). The availability, effectiveness, and access to antenatal care are linked with good perinatal outcomes, making it an important determinant in the health of a whole society(1).

The World Health Organization (WHO) urges countries to expand their agendas to look beyond survival, maximising the health and potential of their populations(5). Best evidence needs to be integrated into practice, whilst certain services should be reconsidered(1). Sustainable and adequate health policies are key to delivering the best possible care to a population, responding adequately to its changing needs(6). Research demonstrates the fundamental aspects of antenatal care, but governments are ultimately responsible for care provision and deciding what aspects are included in the service they provide(7). For the purposes of this review, antenatal care is all the care that a pregnant woman receives from organized health services(1) and antenatal care policy the guidance that aims to draw recommendations on the complex nature of the issues surrounding pregnancy, healthcare practices, and provision(5).

Antenatal care varies within and between countries, sometimes even inside a maternity care setting, in ways that are not fully related to clinical needs, raising questions about the assimilation of evidence into clinical decisions(8), and the existence of clear and updated guidance in the field. In Europe and other high-income countries, perinatal health disparities highpoint both to the need and opportunity for improvement(9). As an example, in the latest European Perinatal report maternal mortality varied from 1.9%_ooo_ to 24.7%_ooo_ (8), episiotomy rates from 4.9% to 75%(10), vaginal birth rates from 39.4% to 77%, and cesarean rates from 16.1% to 56.9%(8). Are these discrepancies related to the organisation of care, and could countries learn from one another?

Previous research explored models of care(11) ideal frequency of antenatal consultations(12) and characteristics of certain care models that may result in improved perinatal outcomes(3). But to the authors’ knowledge, no other review has focused on mapping antenatal care policy, at a country level. To fill this gap, and to feed into further research, the purpose of this scoping review was to identify the antenatal care policies and their characteristics, for low-risk pregnant women in high-income countries with a health care system founded on the Beveridge Model, a universal healthcare system. All evidence gathered will be used by the authors in a cost-effectiveness study comparing a general practitioner-led model of care and a hypothetical midwife-led care model for antenatal service provision in one country with these characteristics: Portugal. The outcomes of this review can inform future policy revision or development in the subject area (e.g.through the identification of gaps in the current policies and differences from best available evidence).

Scoping review was the chosen methodology as it is the most appropriate type of review to identify and map evidence such as policy(13), or simply to identify key characteristics or factors related to a concept(14).

### Review question(s)

What are the antenatal care policies for low-risk pregnant women in high-income countries with a health care system founded on the Beveridge Model?

Additionally, the review addressed the following questions:

i. What are the characteristics of the antenatal care package for low-risk women in each country?
ii. How is the care organized for low-risk pregnant women in each country?
iii. Who provides care for low-risk pregnant women in each country?
iv. What evidence was used to inform the guidance in the field for low-risk pregnant women in each country?

### Inclusion criteria

This review considered documents that included policy or official guidance on antenatal care for low-risk pregnant women in high-income countries, with health care systems comparable to Portugal: Australia, Denmark, Finland, Greece, Iceland, Ireland, Italy, New Zealand, Norway, Portugal, Spain, Sweden, and the United Kingdom (UK).

## Methods

This study was conducted following the JBI methodology for scoping reviews(14) and reported following the PRISMA-ScR guidance(15). An *a priori* protocol(16) has been developed, registered (osf.io/h7um6), and is publicly available. The protocol was methodically followed, and the only change was the removal of “study method” item from the data collection chart.

### Search strategy

Documents published in all languages from 2005 were searched in the main databases such as CINAHL Plus, Scopus, MEDLINE (PubMed), amongst others(16), on March 28^th,^ 2020. Reference lists of the articles selected for full-text review were screened for additional papers and a hand search of grey literature was conducted. Finally, field experts (academics and departments of health) were contacted.

All identified records were collated and uploaded into Mendeley v.1803 and duplicates removed. Two reviewers screened through the records (Figure1).

**Figure 1:**
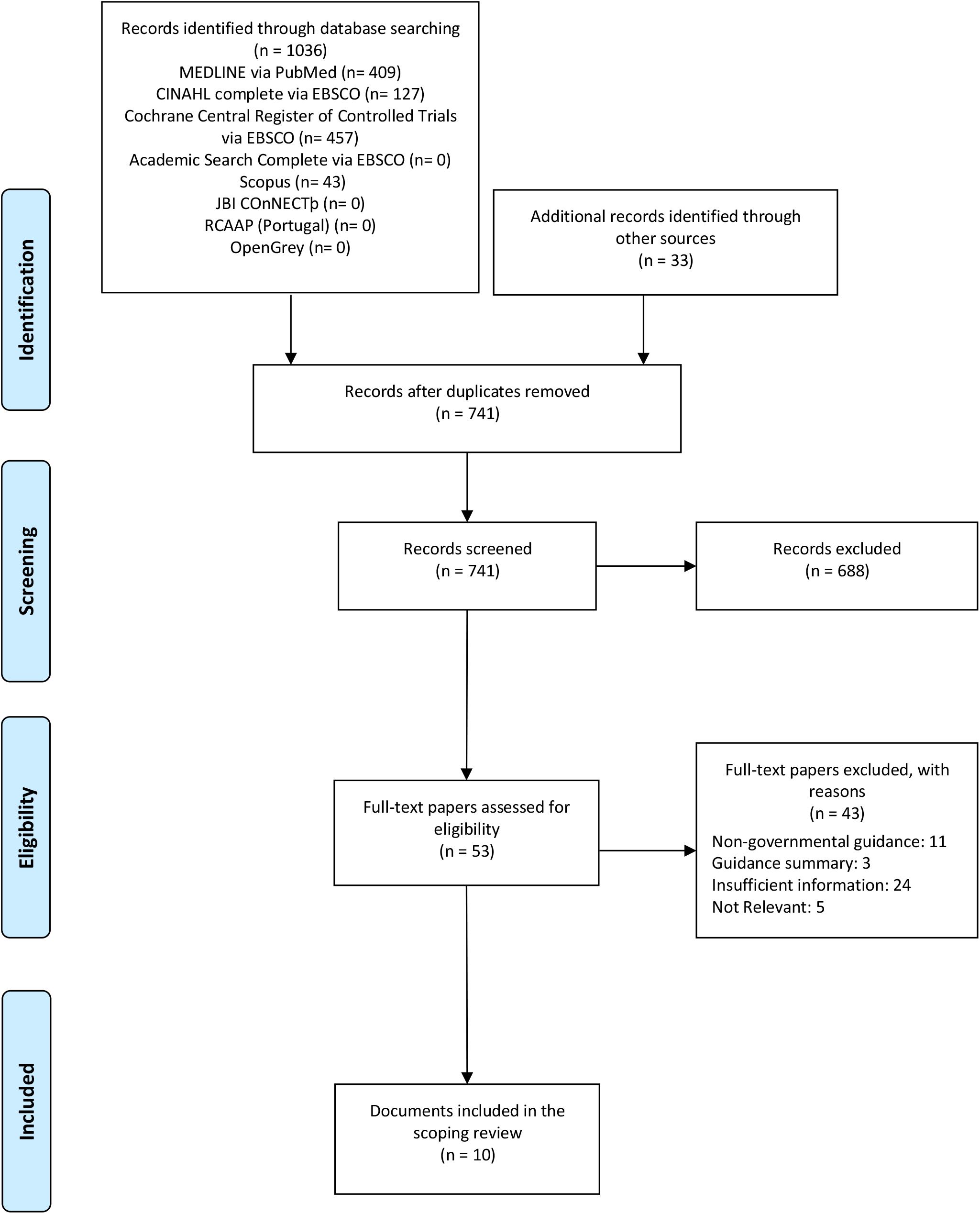
Flow diagram of literature search, study selection, and inclusion/exclusion process, modified from PRISMA(17)

Data was then extracted using a tool(16) previously developed and piloted by the reviewers (tables 2 to 7). The authors did not assess for quality of the documents, since it is not a purpose of a scoping review, but looked at the evidence utilised to inform guidance.

**Table 1.** Included Documents

| Country | Author | Published/Last Updated |
| --- | --- | --- |
| Australia | Australian Department of Health(19) | 2019 |
| Denmark | Danish Department of Health(20) | 2013 |
| Finland | Finnish Department of Health and Welfare(21) | 2013 |
| Iceland | Icelandic Department of Health(22) | 2010 |
| Italy | Italian Ministry of Health(23) | 2013 |
| Norway | Norwegian Department of Health(24) | 2019 |
| Portugal | Portuguese Department of Health(25) | 2015 |
| Spain | Spanish Ministry of Health, Social Services and Equality(26) | 2014 |
| Sweden | Swedish Association for Obstetrics and Gynaecology(27) | 2016 |
| United Kingdom | National Institute for Health and Care Excellence(28) | 2019 |

**Table 2.** Clinical Assessment

| Clinical Assessment | Australia | Denmark | Finland | Iceland | Italy | Norway | Portugal | Spain | Sweden | United Kingdom |
| --- | --- | --- | --- | --- | --- | --- | --- | --- | --- | --- |
| Detailed History | 1<sup>st</sup> | 1<sup>st</sup> | 1<sup>st</sup> | 1<sup>st</sup> | 1<sup>st</sup> | 1<sup>st</sup> | 1<sup>st</sup> | 1<sup>st</sup> | 1<sup>st</sup> | 1<sup>st</sup> |
| Body Mass Index | 1<sup>st</sup> | 1<sup>st</sup> | 1<sup>st</sup> | 1<sup>st</sup> | 1<sup>st</sup> | 1<sup>st</sup> | 1<sup>st</sup> | 1<sup>st</sup> | 1<sup>st</sup> | 1<sup>st</sup> |
| Weight | 1<sup>st</sup> | 1<sup>st</sup> | e.c. | 1<sup>st</sup> | 1<sup>st</sup> | E.c. | E.c. | 1<sup>st</sup> | 10-12w +<br>24-25w +<br>35-36w | 1<sup>st</sup> |
| Blood Pressure | e.c. | e.c. | e.c. | e.c. | e.c. | e.c. | e.c. | e.c. | e.c. | e.c. |
| Urine Reagent Strips | 1<sup>st</sup> | e.c. | e.c. | e.c. | Not clear<br>if routine<br>check only<br>at 1st or at<br>e.c. | e.c. | e.c. | e.c. | e.c. | e.c. |
| Symphysis Fundal Height | e.c. >=24w | e.c. >=24w | e.c. >=24w | e.c. >=25w | e.c. >=28w | e.c. >=24w | e.c. >=14w | e.c. >=24w | e.c. >=24w | e.c. >=24w |
| Abdominal Examination to Identify Fetal Position | e.c. >=36w | e.c. >=36w | e.c. >=30w | e.c. >=36w | e.c. >=36w | a.c. >=36w | e.c. >=36w | e.c. >=36w | e.c. >=35w | e.c. >=36w |
| Routine Fetal Heart Rate Auscultation | Optional | Optional | e.c. >=13-18w | No | Does not<br>provide<br>recommen-<br>dation | e.c. >=24w | e.c. >=12w | Does not<br>provide<br>recommen-<br>dation | e.c. >=24w | Optional |

1<sup>st</sup>: first consultation; w: weeks' gestation; e.c.: each consultation

This review is a secondary analysis of publicly accessible documents and therefore exempt from ethical approval(18).

## Results and Discussion

The search identified 1036 records in the databases, and an additional 33 were found through other sources. After the removal of 328 duplicates and exclusions for several reasons (Figure 1) a total of 10 documents were included in this scoping review.

### Characteristics of the included studies

The review identified the antenatal care policies for all countries except for Greece, the Republic of Ireland, and New Zealand.

### What are the characteristics of the antenatal care package for low-risk women in each country?

The authors looked at routine clinical assessment and antenatal screening (tables 2-4).

**Table 3.** Antenatal Screening: Ultrasound Scans and Chromosomal Anomaly Screening

| Antenatal Screening | Australia | Denmark | Finland | Iceland | Italy | Norway | Portugal | Spain | Sweden | United Kingdom |
| --- | --- | --- | --- | --- | --- | --- | --- | --- | --- | --- |
| **Ultrasound Scans** |  |  |  |  |  |  |  |  |  |  |
| First Trimester | 8-13+6w | 11-13+6w | 10-13+6w | No | 11-13+6w | No | 11-13+6w | 11-13+6w | 10-13+6w | 10-13+6w |
| Second Trimester | 18-20w | 18w | 18w-22w | 19-20w | 19 - 21w | 17-19w | 20-22+6w | 18-22w | 18-20w | 18-20+6w |
| Third Trimester | No | No | No | No | No | No | 30-32+6w | No | No | No |
| **Chromosomal Anomaly Screening** |  |  |  |  |  |  |  |  |  |  |
| Combined 1st trimester screening (MA + NT + free β-hCG + PAPP-A) | 11-13w+6 | 8-13+6w | 11-13+6w | No | 11-13+6w | No | 11-13+6w | 11-13+6w | 10-13+6 | 11-13+6w |

w: weeks' gestation; MA: Maternal age; NT: Nuchal translucency measurement; free $\beta$ -hCG: serum-free beta component of human chorionic gonadotrophin; PAPP-A: Pregnancy-associated plasma protein A; AFP: Alpha-fetoprotein; uE3: unconjugated oestriol.

**Table 4.**
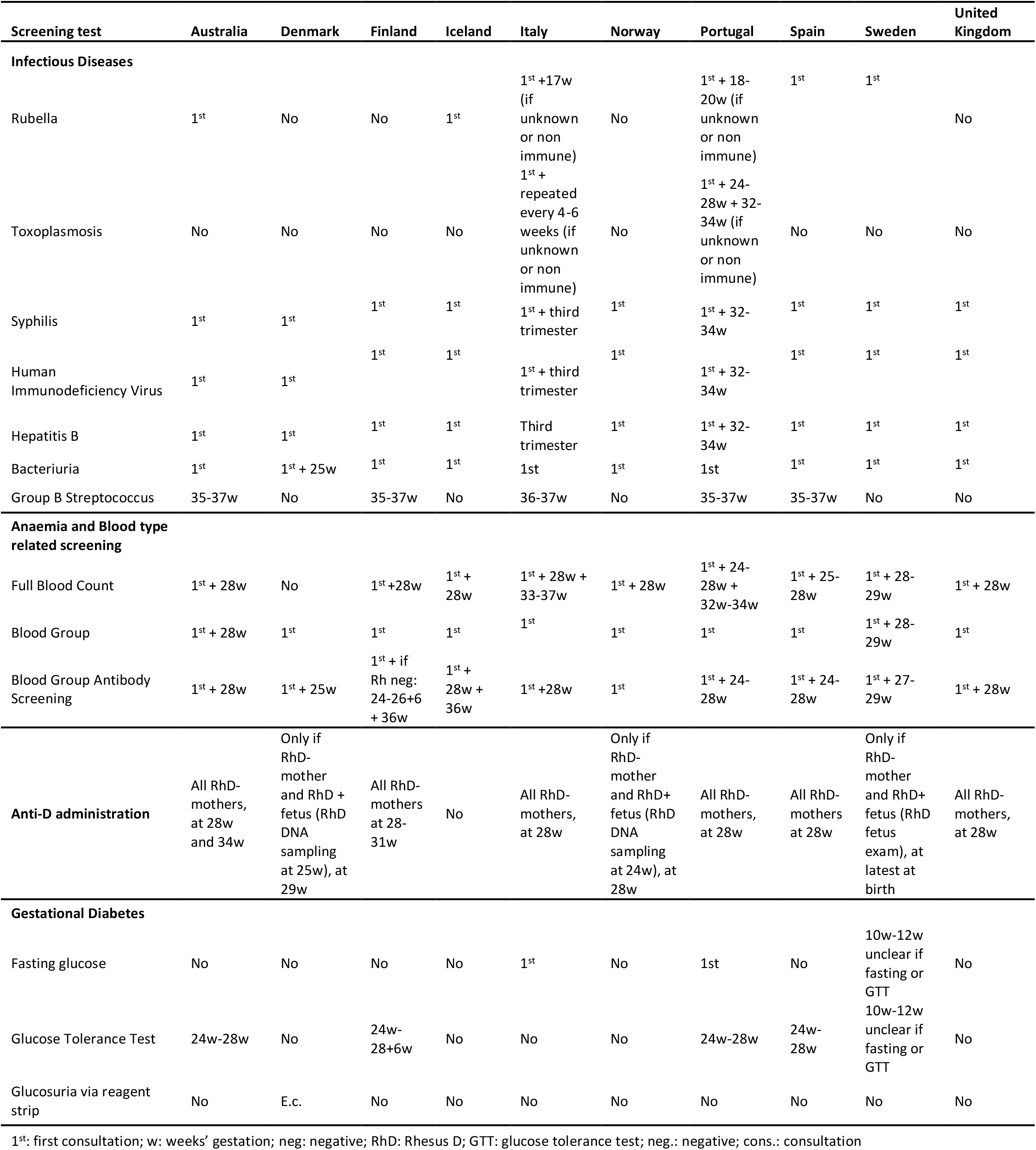
Antenatal Screening: routine urine and blood tests

The clinical assessment presents many aspects of consensus, especially where evidence is strong. The authors found differences in recommendations where evidence seems to be debatable or where practices have been long-standing yet current evidence shows differently. This is the case of routine weight measurements where some countries recommend it throughout pregnancy whilst others encourage self-monitoring (Australia), or no weight checks past the first consultation. In fact, there is no clear evidence that weight measurement has the potential to change maternal and fetal outcomes though it is well established that excessive weight gain during pregnancy is linked to negative outcomes(29). Similarly, routine urine strip tests are only done if risk factors are identified in Australia and Italy, whilst all other countries recommend both blood pressure monitoring and urine strip testing in all consultations. These have historically been conducted routinely aiming to detect pre-eclampsia. Yet, new evidence has found that urine strip testing is inaccurate in predicting significant proteinuria(30) and some experts defend that without risk factors it is unnecessary since it is of little or no benefit in predicting pre-eclampsia(31). Likewise, the amount of proteinuria does not seem to be related to poor maternal and neonatal outcomes(32).

There is expert agreement that blood pressure monitoring is an important intervention in all antenatal care consultations, and the most important factor that influences maternal and neonatal outcomes in the case of pre-eclampsia or hypertensive disorder(32).

Lastly, though evidence is clear that intermittent fetal heart rate monitoring during pregnancy has no predictive value on the pregnancy outcome(33), many countries still recommend it routinely. Finland sustains that listening to the fetal heart rate during normal pregnancy is likely to be important to the woman and her family, which is also the reason why Australia, Denmark, and the UK recommend it as an “optional” intervention. Italy makes no recommendation and Iceland acknowledges its long tradition, though does not recommend it.

Uncommon routine aspects of care (e.g., abdominal circumference measurement - Portugal) were not addressed in this review.

Wide variations in the recommendations for antenatal screening through ultrasound scans (USS) were found, both in frequency and timing. Norway and Iceland’s policies only recommend one USS which is in line with WHO(34) guidance. This scan aims to detect multiple pregnancy and fetal abnormalities, estimate gestational age, and improve a woman’s pregnancy experience(5). All other countries advise an additional USS, where combined screening is offered, generally between 11-13+6 weeks gestation(35).

Ultrasound scanning is considered one of the most important advances in Obstetrics in the 20th century(36) yet its performance is not without risk; such as misdiagnosis/relevance of findings and the risk of possible undesired effects(37).

Portugal is the only country that recommends a routine third trimester USS. The other countries only recommend it based on need. In fact, evidence is in favour of its selective use since in low-risk pregnancies this intervention did not prove to reduce the incidence of adverse perinatal outcomes compared to the selective cases approach(38).

There is consensus in many of the investigations recommended throughout pregnancy however, once again, some areas present wide differences.

Denmark does not recommend a full blood count at the first antenatal check; instead, the policy recommends to universally supplement every pregnant woman with iron, a recommendation not shared by any of the other countries. Italy and Portugal additionally screen for anaemia around 32-37 weeks gestation, which is in line with WHO(5) recommendations, since fetal demands of iron increase significantly in this period(39). Yet, there is a lack of evidence that routine screening for anaemia in asymptomatic women is necessary(40).

Blood group determination is repeated in Australia and Sweden early into the third trimester, but both policies do not provide a rationale for the recommendation. Regarding blood group antibody screening, both Finland and Iceland recommend screening three times during pregnancy, instead of the two times advised by the other countries. However, for both these countries, once the blood group and Rh-D status are determined, repeating antibody screening is only offered to rhesus-negative women.

Screening for Toxoplasmosis is not recommended in any country except for Portugal and Italy. Italy justifies that the pertinence of the recommendation is due to the high incidence of seronegative pregnant women and Portugal does not provide a rationale. The remaining countries advise prevention and education. There is a lack of evidence that antenatal screening and treatment reduces mother-to-child transmission or infection complications(41) and some authors agree that screening has the potential to do more harm than good(42).

Denmark, Finland, Norway, and the UK do not recommend rubella screening. They base their guidance on the premise that screening does not give any protection to the unborn baby(43) and being fully immunised before becoming pregnant is the most effective way to protect women against rubella in pregnancy.

Group B *Streptococcus* (GBS) is one of the tests that often creates divisive opinions. Half of the countries recommend the vaginal/anal swab test whilst the other half do not. Evidence about the benefits of universal screening is limited. Studies have identified reductions in the incidence of early-onset infected newborns, born to mothers identified positive through routine/risk-based antenatal testing and treated with antibiotics in the intrapartum period(44). On the other hand, no differences were found for late-onset of infections. Other studies highlight that infected infants are often born to Group B *Streptococcus* culture-negative mothers and only very few culture-positive mothers will infect their babies(44). Concurrently there is a debate over the exposure to antibiotics and whether the risk of potentially harmful effects is counterbalanced, or not, by the reduction in the incidence of neonatal and maternal sepsis(44).

Finally, screening for Gestational *Diabetes Mellitus* is risk-based in Iceland, Norway, and the UK, and universal in the remaining countries. Variability is visible in the type of test used. Danish policy advises that glucosuria should always trigger a glucose tolerance test. Norway uses HbA1c test in risk-based screening in the first trimester. This variability of approaches is a mirror of the lack of clear evidence. While Gestational *Diabetes Mellitus* is a condition with a considerable prevalence, there is no universally accepted test or diagnosis regimen. Evidence also demonstrates that although gestational diabetes is more likely to be detected when all women are tested, the effects of subsequent management on health outcomes are unclear(45).

Uncommon routine tests (e.g., thyroid function – Spain, hepatitis C – Australia, hemoglobinopathies -UK and Italy) were not addressed in this review.

### How is the care organized for low-risk pregnant women in each country?

Regarding the organisation of care, we looked at the schedule recommended by each country which included both the number of recommended consultations and timings (table 5).

**Table 5.**
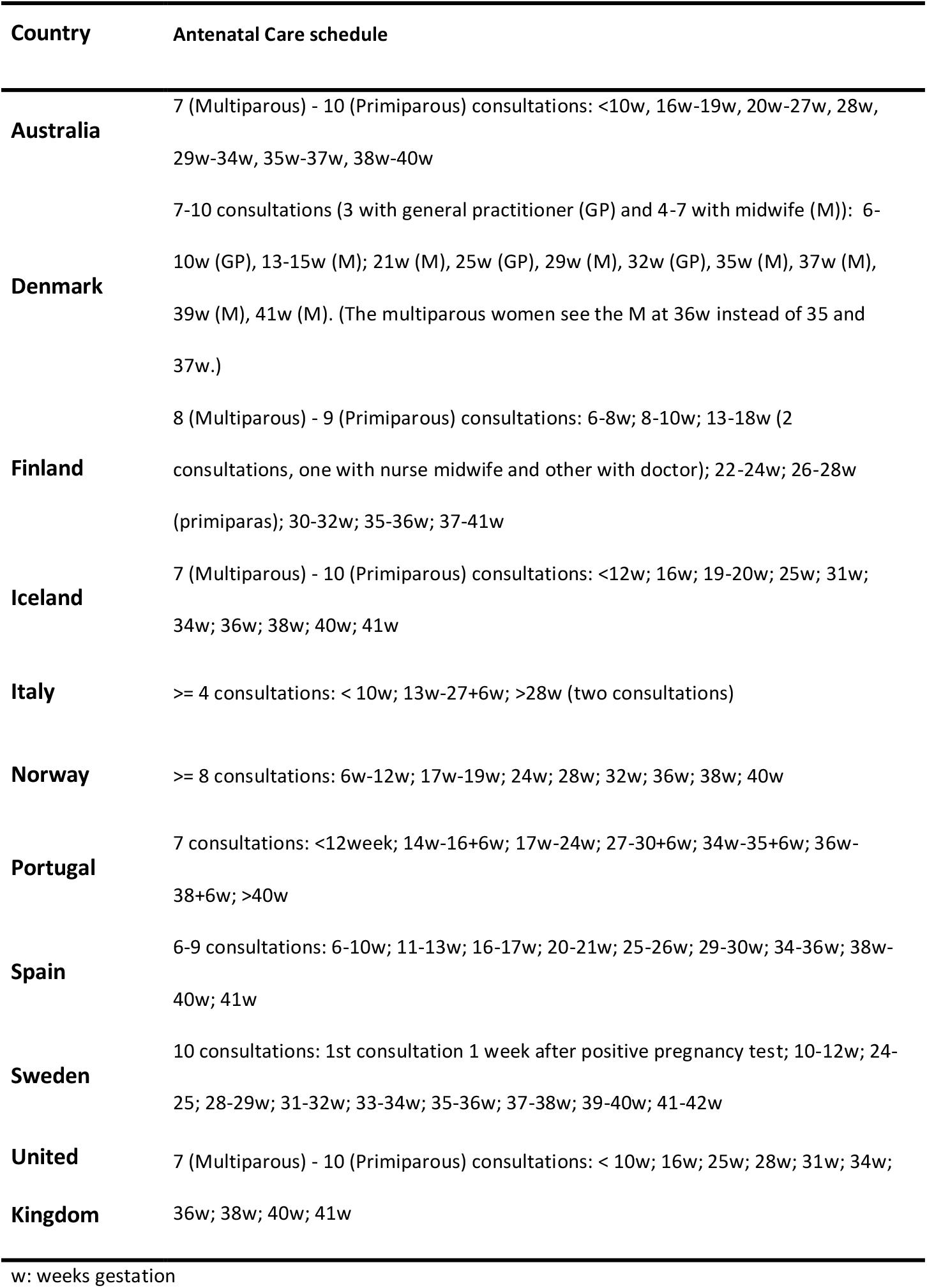
Antenatal care schedule

Regarding the schedule of care, the results demonstrate wide variation. Half of the countries recommend a different frequency of appointments for multiparas and primiparas; the other half recommends the same frequency. None give a clear justificiation for the recommended frequency, although NICE guidance cites a study where women over 35 years of age with previous pregnancies (amongst others characteristics) preferred fewer appointments (46). All make the reservation that the schedule of consultations should always be determined according to the woman’s individual needs.

There is inconclusive evidence as to the “ideal” number of consultations; however, in 2016 the WHO doubled the recommended minimum number of consultations, from 4 to 8 (5). This was based on the probable association of the 4 consultations schedule with more perinatal deaths and evidence supporting the improvement of safety during pregnancy through increased frequency of maternal and fetal assessments to detect problems(5). Evidence also indicates that more contact between pregnant women and a knowledgeable, supportive and respectful antenatal care provider is likely to result in greater maternal satisfaction and a positive pregnancy experience(47). Nonetheless, studies from high-income countries, comparing models with minimum 8 consultations and models with 11-15 consultations, indicate no important differences in maternal and perinatal outcomes, making the earlier more cost-effective(12).

Italy and Portugal do not meet the minimum WHO recommended frequency of consultations (figure 2). This may happen because the latest WHO recommendation was published after the Italian and Portuguese policies (2011 and 2015, respectively).

**Figure 2.**
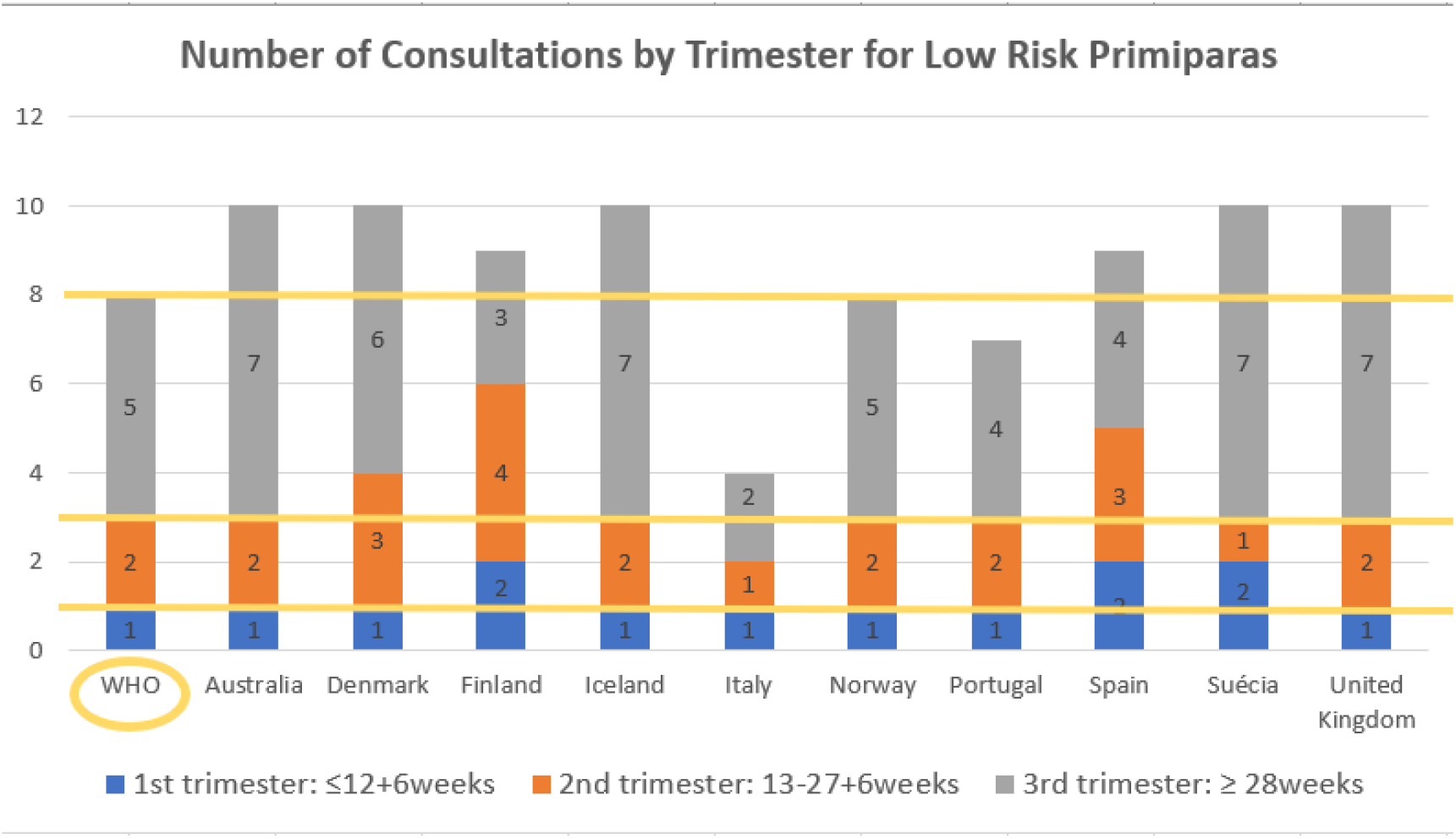
Number of recommended consultations for low-risk primiparas by country and WHO

The timing of the appointments also varies extensively. The most recent WHO recommendation(34) proposes one consultation in the first trimester, two in the second, and five in the third. Neither WHO(5) or the policies present a rationale for the exact gestation they recommend each consultation.

### Who provides care for low-risk pregnant women in each country?

The authors looked at both the recommendation of professional that provides care and the model of care (table 6).

**Table 6.** Recommended main professional that provides care and model of care

| Country | Main professional that provides care | Model of care |
| --- | --- | --- |
| **Australia** | Mw, GP or/and OB | Specific recommendation for continuity of carer |
| **Denmark** | Mw and GP | Specific recommendations for continuity of carer |
| **Finland** | Mw and D | Specific recommendation for continuity of carer |
| **Iceland** | Mw and GP | Specific recommendations for continuity of carer |
| **Italy** | Mw, GP or/and OB | Specific recommendation for continuity of carer |
| **Norway** | Mw and/or D | Specific recommendation for continuity of carer |
| **Portugal** | Unclear. Along the document refers to D. | No recommendation |
| **Spain** | Mw or Mw and GP | No recommendation |
| **Sweden** | Mw | Specific recommendation for continuity of carer |
| **United Kingdom** | Mw or GP | Specific recommendations for continuity of carer |

Mw: Midwives/Nurse Midwives; GP: General Practitioner; OB: Obstetrician; D: Unspecified Doctor

The majority of the policies recommend midwives/nurse-midwives for this role under a continuity of carer model. In fact, the best available evidence supports this recommendation and has consistently demonstrated that women cared under this model are less likely to experience intervention, and more likely to experience positive outcomes(11).

The only two countries that do not propose a continuity of carer model are Spain and Portugal. The latter does not specify the professionals responsible for the provision of antenatal care, although along the document the “doctor” is occasionally mentioned and midwives/nurse-midwives are never referred to.

Despite acknowledging that the midwife-led continuity model of care is the model that results in better outcomes for low-risk pregnant women, many countries also include shared care models with the general practitioner/unspecified doctor or general practitioner-led care. Italy and Australia are the only policies to also acknowledge obstetricians for this role. Evidence demonstrates that routine involvement of obstetricians in the care of women with uncomplicated pregnancies at scheduled times does not appear to improve perinatal outcomes compared with involving obstetricians when complications arise(48).

### What evidence was used to inform the guidance in the field for low-risk pregnant women in each country?

The evidence referred to in guidance as having informed it is summarised in table 7.

**Table 7.** Evidence used to inform the guidance

|  | Literature Search Strategy | Levels of evidence used to support recommendations. |
| --- | --- | --- |
| **Australia** | For all the subject areas, a comprehensive literature search was conducted. References after each section. | Levels of evidence were considered and the highest levels of evidence used. |
| **Denmark** | Does not mention the strategy for the literature search. References in a separate document. | No mention of whether levels of evidence were considered. Policy currently being updated. |
| **Finland** | For all the subject areas, a comprehensive literature search was conducted. References after each section. | Levels of evidence were considered and the highest levels of evidence used. |
| **Iceland** | The policy is based on NICE guidance for antenatal care 2008 and adapted to the national context. Does not present reference lists. There are some hyperlinks along with the document, pointing to places where references can be found, but not directly to the specific issues. | NICE considers levels of evidence for its recommendations. However the Icelandic policy is not in line with the two latest NICE updates. |
| **Italy** | The policy is based on NICE guidance for antenatal care 2008 and adapted to the national context. References after each section. | NICE considers levels of evidence for its recommendations. However the Italian policy is not in line with the two latest NICE updates. |
| **Norway** | For all the subject areas, a comprehensive literature search was conducted, and levels of evidence were established. References after each section. | Levels of evidence were considered and the highest levels of evidence used. |
| **Portugal** | Does not mention the strategy for the literature search. References at the end of the document. | No mention of whether levels of evidence were considered. |
| **Spain** | For all the subject areas, a comprehensive literature search was conducted. References at the end of the document. | Levels of evidence were considered and the highest levels of evidence used. |
| **Sweden** | Does not mention the search strategy for the literature search. References after each section. | No mention of whether levels of evidence were considered. |
| **United Kingdom** | For all the subject areas, a comprehensive literature search was conducted. References at the end of the document. | Levels of evidence were considered and the highest levels of evidence used. |

The results show that all countries provide a degree of evidence for their recommendations. Most present a comprehensive literature search, where levels of evidence were established, and the highest levels of evidence used to support the recommendations. Iceland and Italy based their guidance on NICE (UK) recommendations, with adaptations to their country context yet their recommendations are currently outdated. Denmark, Portugal, and Sweden do not mention their search strategy although they provide partial references for their recommendations.

The absence of a clear strategy to use evidence to inform guidance, as well as the use of evidence that currently is outdated, or is not the best available, demonstrates the need for the policies to be updated. Though the best available research-derived evidence is a key feature of most policy models(49), it is known that often this does not happen due to conflicts that can arise, unrelated to research, though inhibiting its use in policymaking(50). Policymakers have to operate on various competing interests(49) which include finance, cultural beliefs, trade-offs, prejudice, agendas promoted by interest groups threatened by new public regulations, amongst others. All this determines whether research evidence is translated to health policy(50) and can be an explanation for the variability in the recommendations.

### Limitations

Since the policies are written in the country’s mother language and although the relevant information was translated into English, the authors felt they could be missing important information or interpreting differently from the intended. This limitation was minimised by asking bilingual experts to double-check and validate the extracted data.

## Conclusions

The analysed policies have areas of consensus amongst their practices, but relevant variations in care provision were identified, that would not be explained by country context differences and can have an impact on perinatal outcomes, pregnancy, and maternity experience, and/or costs. Some recommendations are not based on the latest best available evidence and need updating.

As previously mentioned, the provision and extent of antenatal care can affect the health and well-being of women and infants. Good outcomes are directly linked with effective and affordable interventions. It is crucial and an ethical necessity that health policies are carefully developed, up to date, and based on the best available evidence, to ensure that all women and babies have the opportunity to achieve the highest standard of health.

Research correlating these results with perinatal outcomes and cost evaluation could be valuable to optimise guidance on antenatal care and consequently health care outcomes. Similarly, some aspects of care (e.g., Gestational *Diabetes Mellitus*, Group B *Streptococcus)* screening, and others, need further rigorous studies to obtain evidence of higher quality to inform recommendations.

## Data Availability

The data referred to in the manuscript is of public access.

## Acknowledgements

I would like to thank Dr Angela Brown and Dr Julie Fleet, Dr Eva Rydhal, Dr Raussi Letho, Dr Laura Batinelli, Dr Fátima Martins, Dr Anna Martin Arribas, and Dr Angela Velinor for their availability to assist with sourcing the correct document and validate the extracted data. This review will contribute towards a PhD in Nursing Sciences for the first author, AG.

